# Perceptions of HPV Self-Collection for Cervical Cancer Screening Among Mobile Health Program Attendees

**DOI:** 10.64898/2026.05.01.26352235

**Authors:** Arturo Tovar, Nicole Person-Rennell, Gloria Coronado, Purnima Madhivanan, Sheila Soto, Haley Escheman, Anna M. Morenz

## Abstract

**Background:** Mobile health programs (MHPs) provide essential preventive services to uninsured and underserved communities. Following the 2024 regulatory approval of human papillomavirus (HPV) self-collection for cervical cancer screening, MHPs represent an access point for healthcare-based self-collection. However, little is known about patient perceptions of this approach in MHP and other healthcare settings.

**Methods:** From May – August 2025, we surveyed individuals aged 25–65 years with a cervix who attended MHPs in Southern Arizona. The survey assessed interest in HPV self-collection, preferred locations, instructional preferences, and facilitators to attend follow-up after a positive result. Descriptive statistics summarized demographic characteristics and survey responses.

**Results:** Fifteen female participants completed the survey (mean age 36 years). Ten (67%) identified as Hispanic or Latino, nine (60%) preferred Spanish, and 14 (93%) were uninsured. Interest in HPV self-collection was high, with ten (67%) very or extremely interested. Among those interested, nine (69%) preferred home-based self-collection, and four (31%) preferred clinic or MHP-based self-collection. Most common concerns regarding self-collection on the MHP were ensuring privacy (n=7; 47%) and knowing how to perform the test correctly (n=5; 33%). Most participants (n=11; 73%) reported being very or extremely confident they would attend follow-up after a positive result; language-concordant support, reminder calls, and scheduling assistance were the most endorsed facilitators.

**Conclusion:** HPV self-collection was highly acceptable among MHP attendees, although home-based self-collection was most preferred. Addressing privacy concerns, providing multiple modes of instruction, and offering navigation support may improve implementation success and ensure timely follow-up care in MHP settings.

## INTRODUCTION

Mobile health programs (MHPs) deliver essential preventive services to uninsured and underserved communities by traveling directly to neighborhoods, reducing logistical barriers, and often providing free care. Currently, approximately 1,300 mobile clinics operate nationwide, serving a population that is 41% uninsured.^1,2^ With anticipated federal cuts to Medicaid and the Affordable Care Act, the uninsured population is projected to grow by 10 million by 2034, further increasing demand for safety-net care including MHPs.^3^

Cancer screening is a key preventive service that MHPs are able to facilitate, alongside vaccinations, infectious disease and blood pressure screening, health education and counseling, and chronic condition diagnosis and management. The most common cancer screening facilitated by MHPs is mobile mammography, and prior literature has demonstrated the ability of MHPs to increase access for under-screened groups.^4^ However, cervical cancer screening is offered less frequently in MHPs despite the evident need in this population. For example, in one study of a Connecticut-based MHP, less than half of the 1,455 clients had ever undergone Pap testing, highlighting the ability of MHPs to reach key populations at risk for not being screened.^5^

The 2024 FDA approval of human papillomavirus (HPV) self-collection for cervical cancer screening presents new opportunities for MHPs to expand access to screening, as current approvals require specimen collection in healthcare settings such as clinics, MHPs, or pharmacies.^6^ While studies demonstrate high acceptability of mailed, at-home HPV self-collection, few have evaluated perceptions of healthcare-based self-collection, particularly in MHP settings.^7–13^ Additionally, concerns about completing follow-up after positive results may limit dissemination of screening programs in mobile health contexts.^4^ This study explored perceptions of healthcare-based HPV self-collection and barriers and facilitators to follow-up among MHP attendees in Southern Arizona.

## METHODS

### Survey Procedures

From May to August 2025, we surveyed individuals aged 25-65 years with a cervix who attended MHPs run by the University of Arizona (UA) Department of Family & Community Medicine or Pima County in Southern Arizona. Established in 1976, the UA MHP cares for individuals without insurance and provides full-spectrum family medicine care, including pre-natal and primary care, at a variety of outreach sites including schools, food banks, and churches. The MHP is equipped with the appropriate space and clinical staff to provide Papanicolaou (Pap) testing for cervical cancer screening. The Pima County MHP offers similar care, including vaccinations, blood pressure screenings, tobacco cessation resources, and comprehensive reproductive health and family planning services at community locations such as county libraries, employment centers, and parks. Using convenience sampling, eligible patients were invited to complete a bilingual (English/Spanish) tablet-based survey after their appointments.

Flyers with a survey link were also posted in the MHPs’ clinical areas. Respondents received a small gift card for their participation. The UA Institutional Review Board reviewed and approved this study.

### Outcomes

Using the COM-B framework (Capability, Opportunity, Motivation-Behavior) as conceptual underpinning, the survey assessed interest in HPV self-collection, preferred locations (using rank-ordering), instructional preferences about how to carry out self-collection, and barriers and facilitators to follow-up after a positive result.^14^ Demographic information and social drivers of health were collected, including housing status, health insurance, transportation access, and neighborhood social vulnerability (measured using the Centers for Disease Control and Prevention’s Social Vulnerability Index [SVI]).^15^

### Analysis

Descriptive statistics were calculated for responses to the survey questions, using mean and standard deviation for continuous variables and counts and percentages for categorical variables. Survey responses were collected using Qualtrics, and analysis was completed in Microsoft Excel (Version 16.103.1). To avoid small cell counts <3, some responses were excluded or combined.

## RESULTS

Fifteen female participants completed the survey (mean age 36 years). Ten (67%) identified as Hispanic or Latino, nine (60%) preferred Spanish, and 14 (93%) were uninsured. Participants lived in neighborhoods with high social vulnerability (mean SVI 0.78, standard deviation 0.13). Most (80%) lacked a primary care provider, citing lack of insurance as the primary barrier (93%). Eleven respondents (73%) had their own car to attend appointments or run errands, and four (27%) borrowed a car, received rides from friends, or used rideshare. The majority (n=13; 87%) denied that a lack of transportation kept them from attending medical appointments.

Most resondents (n=13; 87%) had previously undergone cervical cancer screening, with 12 (80%) screened within the past five years. Among those previously screened, four (31%) received their last screening at an ob/gyn clinic, three (23%) at a MHP, and three (23%) at a primary care clinic. Other screening locations included the health department, emergency room, and outside the U.S.

Interest in HPV self-collection was high, with 10 participants (67%) reporting being very or extremely interested (Table 1). Among those slightly to very interested (n=13), at-home self-collection was most preferred (n=9, 69%), followed by clinic- or MHP-based collection (n=4, 31%). Figure 1 demonstrates the preferences for home-, clinic-, MHP-, and pharmacy-based self-collection screening based on the rank-order question from most to least preferred. Home was most frequently ranked first (most preferred), while pharmacies were consistently least preferred. The most common concerns regarding MHP-based self-collection were ensuring privacy (n=7, 47%) and knowing how to perform the test correctly (n=5, 33%). Participants expressed diverse preferences for instructional modalities, including written pamphlets with photos (33%), live in-person explanation (20%), written instructions only (20%), and video instruction (20%).

**Table 1.** Preferences and Concerns Regarding HPV Self-Collection and Follow-Up Care.

| Survey Domain | Response Rate, N (%) |
| --- | --- |
| Interested in HPV self-collection* |  |
| ...Very or extremely interested | 10 (67%) |
| ...Slightly or moderately interested | 3 (20%) |
| Most preferred location for HPV self-collection (out of n=13 interested) |  |
| ...At home | 9 (69%) |
| ...In clinic or in MHP | 4 (31%) |
| Concerns regarding HPV self-collection on MHP |  |
| ...Making sure the result gets back to my regular doctor, if my regular doctor is outside of the MHP | 4 (27%) |
| ...Ensuring privacy | 7 (47%) |
| ...Knowing how to do the self-collection properly | 5 (33%) |
| ...Childcare while I'm doing self-collection, or having enough time to do the self-collection* | 3 (20%) |
| Preferred instruction modality prior to self-collection |  |
| ...Live, in-person explanation with a clinical staff member | 3 (20%) |
| ...A pamphlet with written instructions and photos of how to do the self-collection | 5 (33%) |
| ...A pamphlet with written instructions only | 3 (20%) |
| ...Brief video instruction | 3 (20%) |
| Confidence about attending follow-up after a positive HPV result |  |
| ...Very or extremely confident | 11 (73%) |
| ...Somewhat or moderately confident | 4 (27%) |
| ...Not at all confident | 0 (0%) |
| Facilitators to attending follow-up after a positive HPV results |  |
| ...Somebody who speaks my language attending with me | 4 (27%) |
| ...Someone calling to remind me | 3 (20%) |
| ...Someone helping me make the appointment, or an appointment on weekends or after 5 pm on weekdays* | 3 (20%) |
| ...A voucher for gas or rideshare to attend | <3 |
| ...A voucher for a childcare provider while attending | <3 |

Notes. HPV = human papillomavirus, MHP = mobile health program. \* indicates where variables were excluded or combined to avoid small cell counts <3; when small cell counts could not be re-calculated from other numbers for multiple-response questions, "<3" was shown instead.

**Figure 1.**
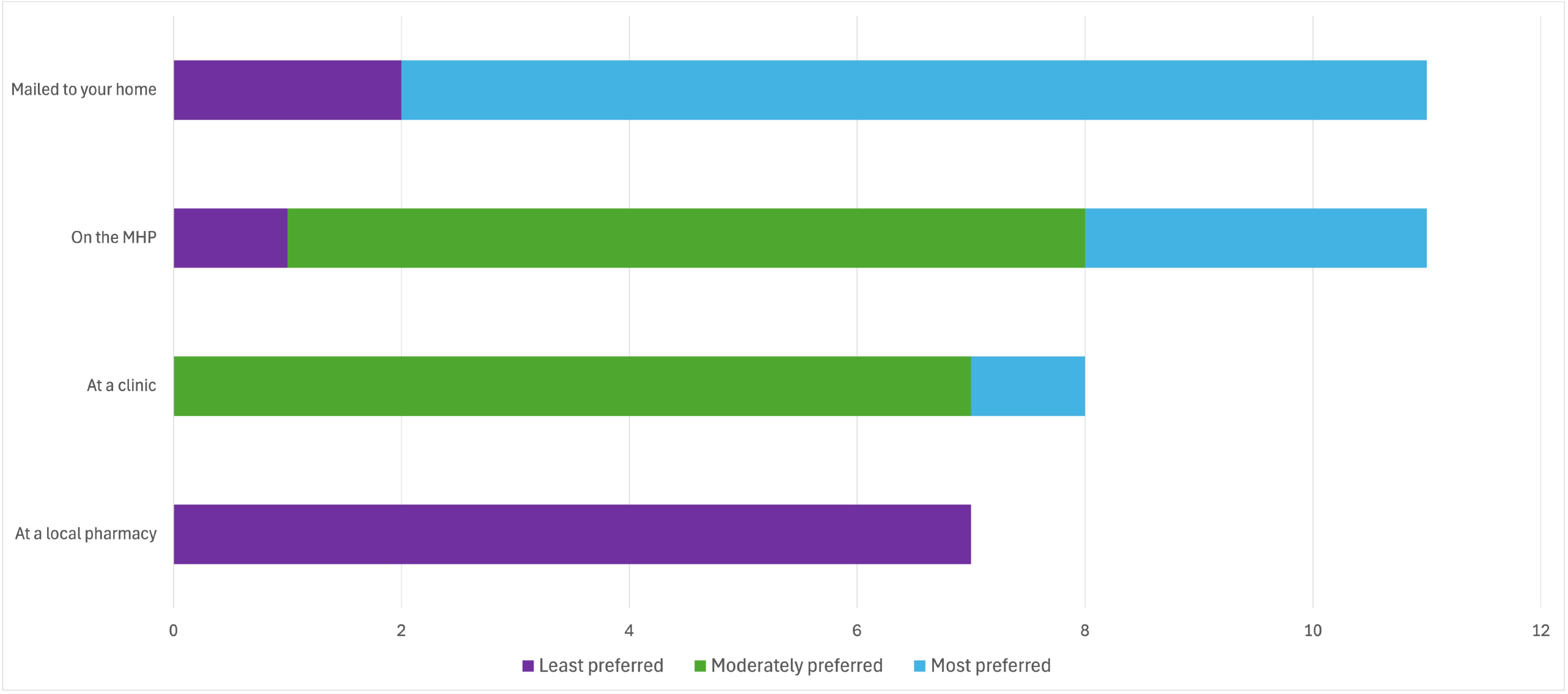
Rank-Order Preferences Regarding Location for HPV Self-Collection Among Mobile Health Program (MHP) Attendees (n=11) Caption: Respondents ranked four potential locations for HPV self-collection (mailed to home, on a mobile health program [MHP], at a clinic, and at a local pharmacy) from most to least preferred. Home was most frequently ranked as most preferred, whereas the pharmacy was consistently ranked as the least preferred location. MHPs and clinics tended to be ranked in the middle (moderately preferred), although they were the most preferred location for some respondents. *Note*. Not all respondents ranked all options available for self-collection locations.

Most participants (n=11, 73%) reported being very or extremely confident they would attend follow-up after a positive HPV result. Key facilitators included having someone who speaks their language accompany them (27%), reminder calls (20%), and scheduling assistance or flexible appointment times on weekends or evenings (20%).

## DISCUSSION

This exploratory study demonstrates high acceptability of HPV self-collection among MHP attendees in Southern Arizona, a predominantly Hispanic, Spanish-speaking, uninsured population with high social vulnerability. Although prior work has demonstrated high acceptability of HPV self-collection, most studies have examined home-based self-collection, while current FDA approval limits the most widely available self-collection kits to healthcare settings.^16–18^ Even though most survey respondents in our study preferred home-based self-collection, nearly one-third preferred clinic- or MHP-based self-collection. This finding may reflect that although clinic-based self-collection has the disadvantages of requiring travel to and added time for a visit, advantages include being able to meet other care needs, receive in-person instruction, and not worry about the sample reaching the lab. The only healthcare location that was consistently not preferred by respondents was the pharmacy, which highlights that while pharmacies are a common setting for health promotion activities, such as vaccination, patients may feel less comfortable with moving cancer screening into this realm and away from the context of longitudinal patient-clinician relationships and familiar clinical settings.

The most common concerns about performing HPV self-collection on the MHP were ensuring privacy and correct self-collection technique. Privacy concerns suggest that MHPs—where clinical space is often limited—may need to adopt deliberate strategies to reassure clients about how their privacy will be safeguarded. Regarding pre-collection instruction, there was no consensus around one preferred modality of instruction, implying that clinics and MHPs may benefit from offering different options or a combination of options depending on client preference and literacy level (such as pamphlets, brief video, and/or in-person explanation).

Despite facing significant structural vulnerabilities, the majority of respondents (80%) were up-to-date on their cervical cancer screening, which differed markedly from the Connecticut-based study finding that less than half of mobile clinic attendees had ever received a Pap test.^5^ This may reflect that many respondents were receiving or had received prenatal and family medicine care on the UA MHP, which could include a Pap test. Nevertheless, respondents who had never received a cervical cancer screening test were very or extremely interested in HPV self-collection, underlining self-collection as an important option to lower barriers to screening and reach never-screened individuals, who face a heightened risk of cervical cancer.^19,20^

Importantly, nearly three-quarters respondents expressed feeling very to extremely confident about following up if they received a positive HPV test result, which is reassuring in the face of concerns that MHP attendees are often a more transient and vulnerable population who may not complete the diagnostic work-up. However, this percentage may be inflated by the connection of many respondents to the UA MHP, where most patients receive a scheduled follow-up appointment prior to leaving the MHP. Respondents stated that follow-up could be enhanced by interventions such as support from a language-concordant navigator or community health worker, scheduling assistance, and flexible appointment times, which should be tested in future work.

Limitations include the small sample size, convenience sampling, and single geographic region, which may limit generalizability. Despite high screening rates in this sample, many MHP populations have lower screening coverage, and findings may differ in settings with less screening history. Last, open-ended survey questions did not receive responses, so further nuances should be explored in greater depth through focus groups or interviews. Nonetheless, these early, exploratory findings provide pragmatic insights into perspectives on healthcare-based HPV self-collection from a high-need, understudied, and mostly uninsured population of MHP attendees. Future work should assess the acceptability, feasibility, equitable delivery, and clinical effectiveness of HPV self-collection delivered in both traditional clinics and MHP settings under current regulatory approval.

In conclusion, HPV self-collection is highly acceptable among MHP attendees, though preferences vary between home- and healthcare-based collection. Successful MHP-based implementation will require attention to privacy and diverse instructional approaches to meet varying health literacy levels and learning styles. Providing language-concordant navigation, scheduling support, and non-traditional appointment times may improve linkages to diagnostic follow-up after a positive HPV result. Larger studies testing HPV self-collection approaches with this high-need population are warranted to expand upon these findings and inform equitable implementation strategies in safety-net and mobile health settings.

## Data Availability

All data produced in the present study are available upon reasonable request to the authors and completion of a data use agreement.

## ACKNOLWEDGEMENTS

Dr. Morenz is supported by the K12 Program at the Southwest Center for Advancing Clinical and Translational Innovation, funded by the National Center for Advancing Translational Sciences under Award Number K12TR005467. The content is solely the responsibility of the authors and does not necessarily represent the official views of the National Institutes of Health.

## REFERENCES

1. Malone NC, Williams MM, Smith Fawzi MC, et al. Mobile health clinics in the United States. Int J Equity Health. 2020;19(1):40. doi:10.1186/s12939-020-1135-7

2. Mobile Health Map. The Family Van. Accessed October 10, 2025. https://www.mobilehealthmap.org/

3. Park E. New CBO Health Coverage Estimates of Budget Reconciliation Law. Georgetown University McCourt School of Public Policy’s Center for Children and Families. August 11, 2025. Accessed December 8, 2025. https://ccf.georgetown.edu/2025/08/14/new-cbo-health-coverage-estimates-of-budget-reconciliation-law/

4. Greenwald ZR, El-Zein M, Bouten S, Ensha H, Vazquez FL, Franco EL. Mobile Screening Units for the Early Detection of Cancer: A Systematic Review. Cancer Epidemiology, Biomarkers & Prevention. 2017;26(12):1679–1694. doi:10.1158/1055-9965.EPI-17-0454

5. Degife EA, Oliveira CR, Znamierowski E, Meyer JP, Sheth SS. Uptake of Cervical Cancer Screening Among Female Patients Using a Mobile Medical Clinic. American Journal of Preventive Medicine. 2023;65(5):835–843. doi:10.1016/j.amepre.2023.05.013

6. FDA Approval of HPV Self-Collection for Cervical Cancer Screening. American Cancer Society. Accessed May 28, 2024. https://pressroom.cancer.org/releases?item=1325

7. Winer RL, Lin J, Tiro JA, et al. Effect of Mailed Human Papillomavirus Test Kits vs Usual Care Reminders on Cervical Cancer Screening Uptake, Precancer Detection, and Treatment: A Randomized Clinical Trial. JAMA Netw Open. 2019;2(11):e1914729. doi:10.1001/jamanetworkopen.2019.14729

8. Winer RL, Lin J, Anderson ML, et al. Strategies to Increase Cervical Cancer Screening With Mailed Human Papillomavirus Self-Sampling Kits: A Randomized Clinical Trial. JAMA. 2023;330(20):1971. doi:10.1001/jama.2023.21471

9. Pretsch PK, Spees LP, Brewer NT, et al. Effect of HPV self-collection kits on cervical cancer screening uptake among under-screened women from low-income US backgrounds (MBMT-3): a phase 3, open-label, randomised controlled trial. The Lancet Public Health. 2023;8(6):e411–e421. doi:10.1016/S2468-2667(23)00076-2

10. Montealegre JR, Hilsenbeck SG, Bulsara S, et al. Self-Collection for Cervical Cancer Screening in a Safety-Net Setting: The PRESTIS Randomized Clinical Trial. JAMA Intern Med. 2025;185(9):1119. doi:10.1001/jamainternmed.2025.2971

11. Aimagambetova G, Atageldiyeva K, Marat A, et al. Comparison of diagnostic accuracy and acceptability of self-sampling devices for human Papillomavirus detection: A systematic review. Preventive Medicine Reports. 2024;38:102590. doi:10.1016/j.pmedr.2024.102590

12. Madhivanan P, Nishimura H, Ravi K, et al. Acceptability and Concordance of Self-Versus Clinician-Sampling for HPV Testing among Rural South Indian Women. Asian Pac J Cancer Prev. 2021;22(3):971–976. doi:10.31557/APJCP.2021.22.3.971

13. Parker SL, Amboree TL, Bulsara S, et al. Self-Sampling for Human Papillomavirus Testing: Acceptability in a U.S. Safety Net Health System. American Journal of Preventive Medicine. 2024;66(3):540–547. doi:10.1016/j.amepre.2023.10.020

14. Michie S, Van Stralen MM, West R. The behaviour change wheel: A new method for characterising and designing behaviour change interventions. Implementation Sci. 2011;6(1):42. doi:10.1186/1748-5908-6-42

15. CDC’s Social Vulnerability Index (SVI). Agency for Toxic Substances and Disease Registry; 2018. Accessed October 18, 2022. https://www.atsdr.cdc.gov/placeandhealth/svi/index.html

16. Cora□Cruz MS, Martinez O, Perez S, Fang CY. Evaluating human papillomavirus (HPV) self□sampling among Latinas in the United States: A systematic review. Cancer Medicine. 2024;13(16):e70098. doi:10.1002/cam4.70098

17. Di Gennaro G, Licata F, Trovato A, Bianco A. Does self-sampling for human papilloma virus testing have the potential to increase cervical cancer screening? An updated meta-analysis of observational studies and randomized clinical trials. Front Public Health. 2022;10:1003461. doi:10.3389/fpubh.2022.1003461

18. Caleia AI, Pires C, Pereira J de F, Pinto-Ribeiro F, Longatto-Filho A. Self-Sampling as a Plausible Alternative to Screen Cervical Cancer Precursor Lesions in a Population with Low Adherence to Screening: A Systematic Review. Acta Cytologica. 2020;64(4):332–343. doi:10.1159/000505121

19. Janerich DT, Hadjimichael O, Schwartz PE, et al. The screening histories of women with invasive cervical cancer. Am J Public Health. 1995;85(6):791–794. doi:10.2105/AJPH.85.6.791

20. Russ S, Kurtz R, Bennett N, Felsen C, Bostick E. Characterization of Cervical Cancer Screening History Among Patients with Invasive Cervical Cancer: A Population-Based Approach. Gynecol Oncol Rep. 2024;55:101480. doi:10.1016/j.gore.2024.101480

